# Standardization of lithium concentration to the 12-hour level using SimpLi: a simulation study and model validation

**DOI:** 10.64898/2026.01.29.26344876

**Authors:** Kasyanov Evgeny Dmitrievich, Mazo Galina Elevna

**Affiliations:** Department of Social Neuropsychiatry of the Federal State Budgetary Institution “V. M. Bekhterev National Medical Research Center for Psychiatry and Neurology” of the Ministry of Health of the Russian Federation; Innovative Scientific Development of the Federal State Budgetary Institution “V. M. Bekhterev National Medical Research Center for Psychiatry and Neurology” of the Ministry of Health of the Russian Federation

**Author notes:** **Corresponding author:** Kasyanov Evgeny Dmitrievich. –. SPIN code: 4818-2523. SPIN code: 1361-6333.

**Keywords:** lithium, therapeutic drug monitoring, bipolar disorder, pharmacokinetic modeling, simulation study, statistical model

## Abstract

**Background:** Lithium is one of the key medications for the treatment of bipolar disorder, but it requires therapeutic drug monitoring because of its narrow therapeutic window. In routine clinical practice, blood sampling is often performed outside the recommended 10–14 hour interval after the last evening dose, which distorts interpretation of the measured concentration (overestimation with early sampling and underestimation with late sampling) and may lead to inappropriate dose adjustment.

**Objective:** To develop and validate, using synthetic data, a multiplicative model (SimpLi) that standardizes a measured lithium concentration to the 12-hour level while accounting for sampling time and daily dose.

**Materials and Methods:** A simulation study was conducted in accordance with ADEMP recommendations. A synthetic cross-sectional dataset (n = 1000) was generated with distributions of time since the last lithium dose, serum concentrations, and doses derived from the Bipolar CHOICE study, with a median sampling time of 12 hours (IQR 11–14) and a time–concentration correlation of r ≈ −0.30. The dataset was split 70/30 with stratification by time intervals, and 5-fold cross-validation was performed. Model performance was evaluated using RMSE, MAE, and R^2^.

**Results:** The simulation closely reproduced the prespecified time distribution, achieved the target time–concentration correlation (r ≈ −0.30), and yielded a clinically plausible dose structure. A model using time as the only predictor showed limited accuracy (RMSE = 0.316; R^2^ = 0.108), while adding dose provided a moderate improvement (RMSE = 0.303; R^2^ = 0.177). When sampling occurred exactly at 12 hours, direct prediction was biased (−0.150; RMSE = 0.357), supporting the need for an individual correction factor. In a proof-of-concept analysis of five clinical cases, SimpLi produced a lower MAE than the eLi_12_ formula (0.042 vs 0.056 mEq/L).

**Conclusions:** SimpLi is a practical tool (psyandneuro.ru/bekhterev-ai/simpli/) for standardizing lithium levels to 12 hours when sampling times vary. External validation on real-world data and robustness testing across clinical scenarios are needed.

## Introduction

Lithium has long remained one of the cornerstone treatments for the prophylaxis and management of bipolar disorder owing to its mood-stabilizing (normothymic) effects [1]. However, it has a narrow therapeutic window, and its effective and safe use therefore requires regular therapeutic drug monitoring of blood lithium concentrations [2]. The standard approach to lithium level monitoring, established more than 55 years ago, is blood sampling 12 hours after the last evening dose [3]. Adherence to this interval is crucial: at concentrations below 0.4 mmol/L, lithium’s efficacy is questionable, whereas above 0.8 mmol/L most patients develop adverse effects, and levels exceeding 1.2 mmol/L carry a risk of intoxication [4]. Nevertheless, most contemporary clinical guidelines accept a wider sampling window of 10 to 14 hours after the last lithium dose [5,6].

In real-world clinical practice, these recommendations are de facto not consistently followed. Recent studies indicate that approximately half of lithium measurements are performed outside the recommended 10–14 hour interval [7]. For example, in the controlled clinical trial Bipolar CHOICE, 44.9% of lithium blood tests were obtained either earlier than 10 hours or later than 14 hours after the last dose [8]. Similarly, in a large real-world practice review, 49.7% of blood draws fell outside the 10–14 hour window [7].

Such deviations from the recommended sampling time have clear clinical implications. Serum lithium concentration is an essential tool for clinicians when titrating the optimal dose, and failure to standardize sampling time can distort results [4]. For instance, if blood is drawn later than 12 hours after the last dose, the measured lithium level will be artificially lower, potentially prompting an unwarranted dose increase and thereby increasing the risk of toxic adverse effects. Conversely, overly early sampling may yield an inflated level, leading to unjustified dose reduction and insufficient treatment efficacy [9].

Thus, standardizing the timing of lithium assays is critical for accurate interpretation of therapeutic monitoring data. Suboptimal adherence to the 12-hour standard in clinical practice underscores the need for measures that improve compliance with this rule. One potential solution is to use machine-learning models to standardize a measured level to the reference 12-hour concentration. The aim of this study was to develop and validate a simple mathematical model (SimpLi) for standardizing lithium concentration to the 12-hour level based on the time elapsed since the last dose.

The choice of a simulation approach as the first stage of this work was driven by several methodological and practical considerations. First, large standardized datasets in Russian clinical practice that include precise information on both lithium dosing time and blood draw time, which are required for initial model development and calibration, are currently unavailable. Second, access to detailed international datasets is often restricted. A simulation study makes it possible, under controlled conditions, to test the concept and feasibility of the algorithm, define different scenarios, and estimate potential model accuracy prior to evaluation in real clinical samples. This approach is consistent with international experience in developing similar tools, where modeling is regarded as a valid and necessary step preceding clinical validation [10].

## Materials and Methods

This study is a simulation investigation conducted in accordance with the ADEMP framework (Aims, Data-generating mechanisms, Estimands, Methods, Performance measures) for reporting and evaluating simulation studies [10]. Following these recommendations, we specified the modeling aims, the synthetic data–generating mechanism, the target estimands, the analytical methods, and the model performance measures in a stepwise manner, thereby ensuring transparency, reproducibility, and interpretability of the results.

### Aims

The analysis was performed in 2026 and aimed to develop and validate statistical approaches for standardizing serum lithium concentrations to the 12-hour level when blood sampling time varies. To this end, we generated a synthetic cross-sectional cohort of patients with bipolar disorder (n = 1000) receiving lithium, with realistic distributions of sampling time relative to the last dose, serum lithium concentration, and daily lithium dose. No individual-level data from the original studies were used.

### Data-generating mechanisms

#### Sources of parameters and target characteristics

Generation parameters were derived from the aggregated descriptive statistics reported by Köhler-Forsberg et al. (2025) [9], calculated from Bipolar CHOICE, a 6-month multicenter pragmatic randomized outpatient trial comparing lithium- and quetiapine-based strategies with flexible, individualized concomitant treatment [8].

Target characteristics included: (i) subgroup sample sizes; (ii) interval-specific means and standard deviations for serum lithium concentration and lithium dose within prespecified time categories (<8, 8–12, 12–14, 14–16, 16–20, >20 hours; the >20 category was interpreted as 20–24 hours); and (iii) the target negative association between concentration and time (Pearson r = −0.30) reported in the source publication.

#### Sample size and allocation across time intervals

The synthetic cohort size was fixed at n = 1000. Allocation to time intervals was set proportionally to the original counts 28/80/39/21/16/8 (total 192) and scaled to n = 1000.

#### Generation of time since last dose (t)

To reproduce the median and interquartile range of sampling time (Me = 12 hours; IQR = 11–14) exactly, individual t values were generated deterministically on an integer-hour grid with prespecified counts per hour within each interval (e.g., <8 corresponded to 0–7; 8–12 included 8–12, etc.). In addition, a “spike” at exactly 12 hours was imposed: the proportion with t = 12 was fixed at 0.276 (53/192) in accordance with the textual description in the source article. Quartiles were computed as empirical quantiles for discrete data (type = 1 in R), ensuring Me = 12 and IQR = 11–14 by construction.

#### Generation of serum lithium concentration

To match the within-interval concentration distributions, we modeled a normally distributed variable *Y* ∼ *N (μ*_*Y*_,*σ*_*Y*_), interpreted as the square root of the concentration, and then back-transformed to *C=Y*^*2*^. The parameters *μ*_*Y*_ and *σ*_*Y*_ were derived analytically so that the mean and standard deviation of *C* on the original scale matched the target values for each interval.

#### Within-interval trend and the β parameter (calibration of the overall time–concentration correlation)

To simulate a continuous decline in concentration with increasing time since the last dose (beyond stepwise differences between intervals), we introduced a linear time effect within each interval on the transformed, centered scale. The magnitude of this within-interval effect was controlled by parameter β, representing the time–concentration slope on the square-root scale in the data-generating model (not a regression estimate from real data). After back-transformation to the original scale, concentrations within each interval were affinely rescaled to restore the target means and standard deviations while preserving the individual time trend. β was calibrated via a one-dimensional root-finding procedure so that the Pearson correlation between concentration and time in the final synthetic cohort equaled −0.30.

#### Generation of daily lithium dose (mg/day) with a prespecified dose–concentration relationship

Dose was generated after concentration and time, with two simultaneous goals: (i) matching the published interval-specific dose means and standard deviations and (ii) inducing a moderate positive association between dose and concentration. We first generated a continuous dose variable linked to concentration using a Gaussian copula approach: concentration values were mapped to a latent normal scale via rank-based transformation, and a latent normal dose variable was then generated with a prespecified correlation with the latent concentration variable. The correlation parameter was selected such that, prior to discretization, the target association was approximately Pearson r ≈ 0.30–0.40, after which the realized correlation was evaluated in the final data. Next, the latent dose variable was transformed to the original dose scale within each time interval via quantile mapping to a target distribution with the published mean and standard deviation. Negative values were truncated as physiologically implausible.

To better reflect clinical practice, dose was additionally discretized to multiples of 150 mg or 300 mg (depending on the chosen step), consistent with available tablet strengths. Discretization was performed via stochastic rounding to neighboring multiples with probabilities proportional to the distance from the continuous dose to the lower and upper multiples, which preserves the dose distribution on average. After rounding, within-interval calibration was applied to ensure close agreement between the discretized dose means and standard deviations and their targets.

### Estimands

At the data-generation stage, the target estimands were the published aggregate characteristics that the synthetic dataset was required to reproduce: (i) the time distribution (median, interquartile range, and the proportion sampled exactly at 12 hours); (ii) interval-specific means and standard deviations of serum lithium concentration and dose; (iii) the concentration–time correlation (Pearson r ≈ −0.30); and (iv) the dose–concentration correlation (target Pearson r ≈ 0.30–0.40 before discretization, followed by verification in the final data).

### Methods

Simulations were implemented in R (version 4.4.0). The MASS package was used to generate multivariate normal variables for the copula step, and dplyr was used for data handling. Calibration of β was performed via one-dimensional root finding using the base stats function *uniroot*. Random-number generation was seeded at key simulation steps to ensure reproducibility.

A single synthetic cohort was generated (number of simulation replicates, nsim = 1). The objective of this stage was to obtain one fixed synthetic dataset meeting the prespecified aggregate targets (time distribution, interval-specific concentration means and standard deviations, and the concentration–time correlation), which was then used as input for modeling and testing the 12-hour standardization algorithms.

To ensure reproducibility, all stochastic components of data generation were run with fixed random seeds. Specifically, a fixed seed (e.g., *set*.*seed(123)*) was used when generating individual concentration and dose values, whereas subsequent procedures that did not affect the generative model itself (row shuffling, train/test splitting, and creation of cross-validation folds) used a separate fixed seed (e.g., *set*.*seed(2026)*) to reproduce the modeling pipeline. Individual time-since-dose values (t) were specified deterministically on an integer-hour grid with fixed hourly counts within intervals, guaranteeing the prespecified quartile characteristics (including median and interquartile range) and the proportion at exactly 12 hours. The full generation code and all R package versions are provided in the appendix/repository.

### Performance measures

Dose simulation validity was assessed by: (i) the proportion of values that were exact multiples of the selected dosing step; (ii) the Pearson correlation between dose and serum lithium concentration; and (iii) discrepancies between target and achieved interval-specific dose means and standard deviations. Concentration and time simulation validity were assessed by agreement in t quartiles and the proportion with t = 12, agreement with target interval-specific concentration means and standard deviations, and attainment of the target concentration–time correlation (Pearson r ≈ −0.30).

### Modeling and statistical analysis

To evaluate generalization performance, data were split into training (70%) and test (30%) sets with stratification by time intervals to preserve identical distributions of time categories in both subsets. On the training set, 5-fold cross-validation was performed, also stratified by time intervals. Predictive performance was evaluated using RMSE (root mean squared error), MAE (mean absolute error), and R^2^ on the original serum lithium concentration scale (mEq/L). Final model generalization performance was assessed on the held-out test set.

Statistical modeling proceeded in three sequential steps:

1. Replication of the original article’s modeling approach. Because serum lithium concentration is skewed, the primary analysis used a Gaussian linear model (ordinary least squares with normally distributed residuals) fitted on the square-root-transformed concentration scale. The outcome was the observed serum lithium concentration (mEq/L), and the predictor was time since the last lithium dose (hours).
2. Model extension by including lithium dose. The outcome remained the observed serum lithium concentration (mEq/L), and predictors included time since last dose (hours) and lithium dose (mg/day).
3. Standardization of serum lithium concentration to 12 hours using the extended model and an individual correction factor that preserves each patient’s deviation from the model’s average trend.

## Results

### Reproduction of the distribution of time since the last lithium dose

As shown in Table 1, the distribution of *t* reproduced the prespecified quartiles exactly (Me = 12 hours; IQR = 11–14). The proportion of observations with *t* = 12 was 0.276, consistent with the target values reported by Köhler-Forsberg et al. (2025) [9].

**Table 1.** Deterministic design of time since the last lithium dose (n = 1000).

| Time interval | Permissible integer hours | Number of participants by hour (t: N) | Total participants |
| --- | --- | --- | --- |
| <8 | 0–7 | 0:3; 1:4; 2:6; 3:9;<br>4:15; 5:22; 6:36;<br>7:51 | 146 |
| 8–12 | 8–12 | 8:7; 9:14; 10:35;<br>11:85; 12:276 | 417 |
| 12–14 | 13–14 | 13:50; 14:153 | 203 |
| 14–16 | 15–16 | 15:76; 16:33 | 109 |
| 16–20 | 17–20 | 17:33; 18:25;<br>19:17; 20:8 | 83 |
| >20 (20–24) | 21–24 | 21:17; 22:13;<br>23:8; 24:4 | 42 |

### Reproduction of descriptive statistics for serum lithium concentration across time intervals

As shown in Table 2, the mean values and standard deviations of serum lithium concentration in the simulated data were virtually identical to the target values across all time intervals. Minor differences on the order of 10^−7^ or smaller were observed only at the level of floating-point machine precision and have no practical relevance. Plots of the lithium distribution in the overall sample and within time intervals are presented in Figure 1.

**Table 2.**
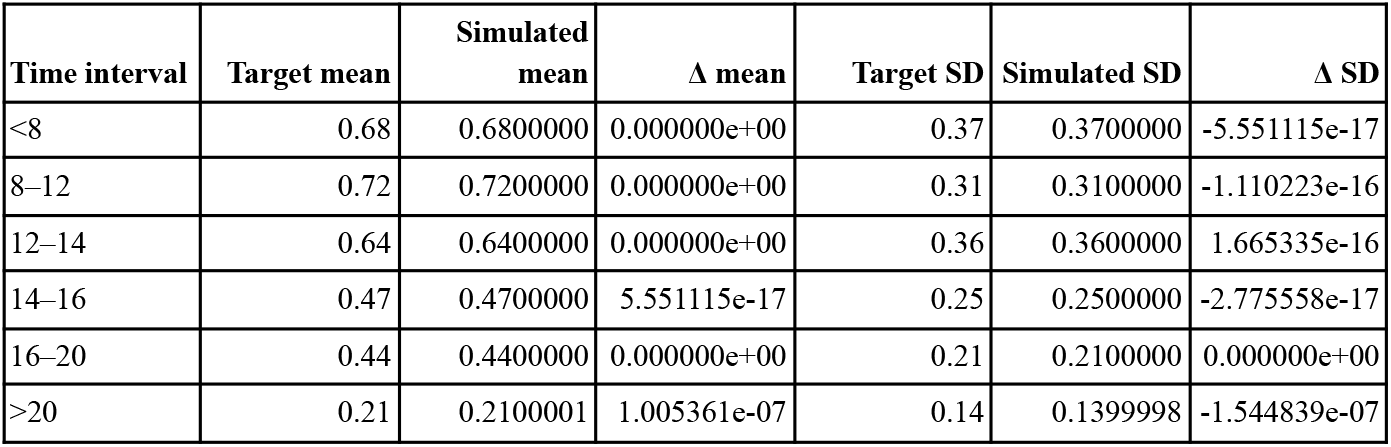
Validation of serum lithium concentration across time intervals.

| Time interval | Target mean | Simulated mean | $\Delta$ mean | Target SD | Simulated SD | $\Delta$ SD |
| --- | --- | --- | --- | --- | --- | --- |
| <8 | 0.68 | 0.6800000 | 0.000000e+00 | 0.37 | 0.3700000 | -5.551115e-17 |
| 8–12 | 0.72 | 0.7200000 | 0.000000e+00 | 0.31 | 0.3100000 | -1.110223e-16 |
| 12–14 | 0.64 | 0.6400000 | 0.000000e+00 | 0.36 | 0.3600000 | 1.665335e-16 |
| 14–16 | 0.47 | 0.4700000 | 5.551115e-17 | 0.25 | 0.2500000 | -2.775558e-17 |
| 16–20 | 0.44 | 0.4400000 | 0.000000e+00 | 0.21 | 0.2100000 | 0.000000e+00 |
| >20 | 0.21 | 0.2100001 | 1.005361e-07 | 0.14 | 0.1399998 | -1.544839e-07 |

**Figure 1.**
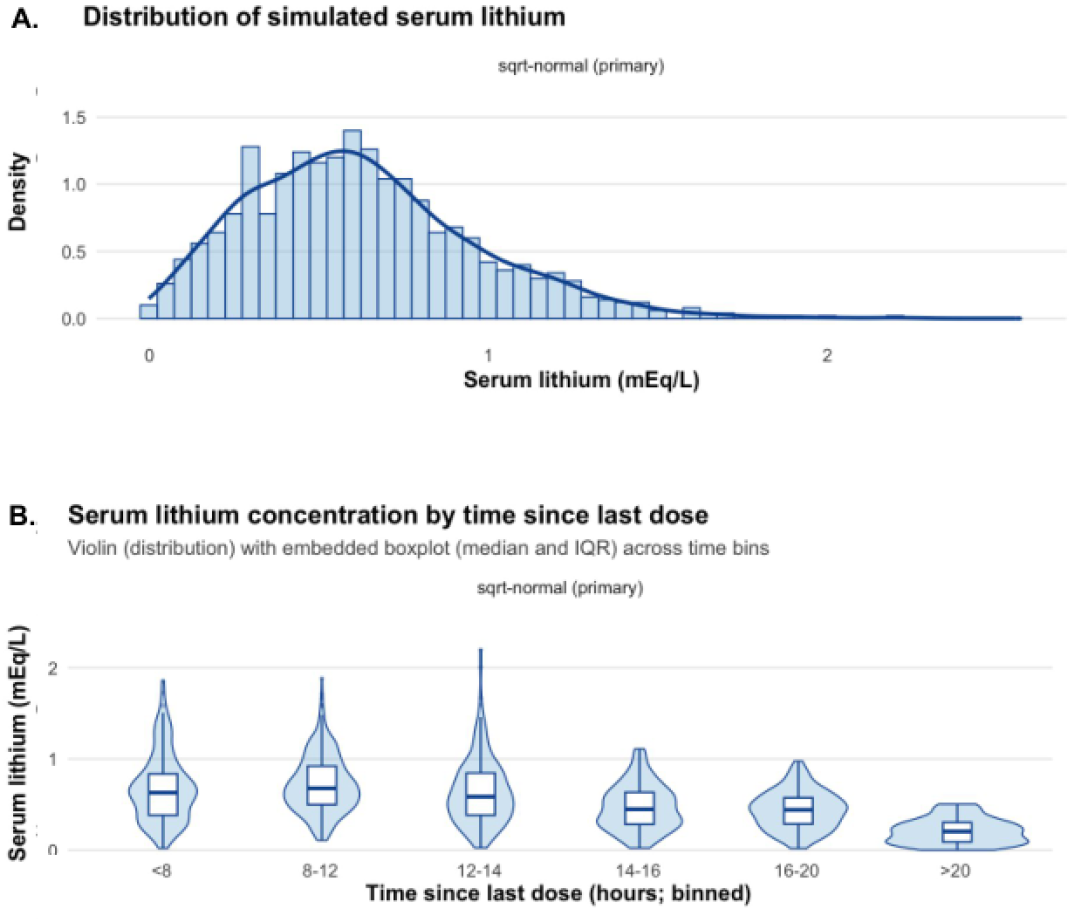
Plots of (A) the distribution of lithium in the overall sample and (B) across time intervals.

### Calibration of the association between serum lithium concentration and time

After tuning parameter β, the target Pearson correlation of r = −0.30 was reproduced with high precision: in the main scenario, Pearson r = −0.2999843, and in the log-normal scenario, Pearson r = −0.3000001. Thus, the simulation simultaneously preserved (i) the interval-specific descriptive statistics of serum lithium concentration and (ii) a continuous negative relationship between serum lithium concentration and time since the last dose.

### Reproduction of descriptive statistics for lithium dose

As shown in Table S3, discretizing the daily lithium dose to values that were multiples of 150 mg and 300 mg produced a fully “clinically plausible” data structure without loss of agreement with the target parameters. In all generated datasets, the proportion of doses that were not multiples of 150 mg or 300 mg was 0, confirming the correctness of the discretization procedure. At the same time, the interval-specific aggregate dose characteristics remained virtually unchanged and matched the published target values: the mean dose in each bin deviated from the specified target by approximately 2 mg in absolute value (the largest observed mean deviation did not exceed ∼1.8 mg), and standard deviations remained close to the targets, with small differences on the order of a few milligrams. Thus, rounding did not distort either the central tendency or the dispersion of dose within the time intervals.

Nevertheless, as expected, discretization attenuated the prespecified positive dose–concentration association compared with the continuous variant: the observed Pearson correlation between dose and serum lithium concentration in the final data was r = 0.33. This reduction reflects the limited “resolution” of tablet-based dosing and is consistent with clinical reality, where even in the presence of an overall dose–response relationship, concentration varies substantially due to interindividual pharmacokinetic differences.

**Table S3.** Verification of lithium dose discretization (150/300 mg step).

| Time interval | Mean dose (simulation), mg | Dose SD (simulation) | Target mean dose, mg | Target SD | $\Delta$ mean dose | $\Delta$ SD |
| --- | --- | --- | --- | --- | --- | --- |
| <8 | 1019.18 | 218.56 | 1021 | 216 | −1.82 | 2.56 |
| 8–12 | 1035.25 | 266.08 | 1034 | 262 | 1.25 | 4.01 |
| 12–14 | 1015.27 | 294.02 | 1017 | 291 | −1.73 | 3.02 |
| 14–16 | 946.79 | 231.57 | 945 | 228 | 1.79 | 3.57 |
| 16–20 | 842.17 | 258.57 | 842 | 256 | 0.17 | 2.57 |
| >20 | 950.00 | 228.73 | 950 | 225 | 0.00 | 3.73 |

### Replication of linear regression results

Linear regression demonstrated a statistically significant negative association between time since the last lithium dose and serum lithium level. The estimated time coefficient was β = −0.0163 (SE = 0.00181; p < 2×10^−16^), corresponding to a decrease of 0.0163 units for each additional hour after dosing. The 95% confidence interval for β ranged from −0.0199 to −0.0128, consistent with the findings of the original article.

In 5-fold cross-validation on the training set, the model achieved RMSE = 0.317 (SD 0.025), MAE = 0.234 (SD 0.015), and a mean R^2^ = 0.041 (SD 0.047). The low R^2^ values indicate that time since dosing explains only a small proportion of the overall variability in serum lithium concentration, which is consistent with substantial interindividual heterogeneity observed in therapeutic drug monitoring.

On the held-out test set, the model achieved RMSE = 0.316, MAE = 0.248, and R^2^ = 0.108. Thus, when using only time since the last dose as a predictor, the linear model showed limited predictive performance but provided a stable and interpretable estimate of the average temporal trend in serum lithium concentration.

### Modeling lithium concentration prediction in the simulated cohort

In the simulated dataset, we fitted a linear regression model in which the outcome was the square-root–transformed serum lithium concentration, and the predictors were time since the last lithium dose (hours) and daily lithium dose (mg/day). To assess generalization performance, the data were split into a training set (70%) and a test set (30%) with stratification by time intervals; 5-fold cross-validation was then performed on the training set. Performance metrics were computed on the original concentration scale (mEq/L) after back-transforming the predictions.

In 5-fold cross-validation on the training set, the mean performance metrics were RMSE = 0.313 (SD 0.021), MAE = 0.240 (SD 0.018), and R^2^ = 0.143 (SD 0.075), indicating stable model performance across repeated training on different subsamples. On the test set, the “time + dose” model achieved RMSE = 0.303, MAE = 0.236, and R^2^ = 0.177, demonstrating limited but reproducible predictive ability. This pattern reflects substantial interindividual variability in therapeutic lithium concentrations even after accounting for dose and time since dosing.

The estimated time coefficient remained statistically significant and negative (β_t = −0.0143), consistent with the expected decline in concentration as time since the last dose increases. The dose coefficient in this specific implementation was small (β_dose = 0.0003), reflecting that within the simulation protocol, dose was introduced as a variable with a prespecified moderate correlation with concentration but without an explicit causal “dose–concentration” mechanism mediated by pharmacokinetic parameters. In addition, part of the dose effect may be “absorbed” by interindividual differences not captured by this simplified model.

### Standardization of lithium concentration to 12 hours using an individual correction factor

We examined patients in the test set whose simulated sampling time was exactly 12 hours (n = 82) and evaluated the model’s direct prediction of concentration. The error was substantial: bias = −0.150, MAE = 0.277, and RMSE = 0.357. This underscores that even when sampling time is correct and dose is known, a large amount of interindividual variability remains unexplained by time and dose. Therefore, an individualized adjustment is biologically plausible.

Accordingly, for the practical task of standardizing a measured concentration to the 12-hour level, we applied a two-step procedure that combines (i) prediction of the “expected” concentration for a given time and dose and (ii) an *individual correction factor* that preserves each patient’s deviation from the model’s average trend.

First, the “time + dose” model is used to compute the expected concentration at the actual blood draw time for the patient’s observed parameters (t_meas and dose); we denote this value as Ĉ_meas. Next, an individual correction factor is calculated as r = C_meas/Ĉ_meas, where C_meas is the measured concentration. This factor can be interpreted as a patient-specific adjustment (“higher/lower than expected”) relative to the sample’s average relationship, and it plausibly captures unmeasured sources of pharmacokinetic variability – such as individual clearance, distribution characteristics, adherence patterns, drug interactions, hydration status, and other factors that are not observed in a simplified model. The same model then computes the expected concentration for the same dose at 12 hours, Ĉ_12. The standardized value is defined as Ĉ_12 = Ĉ_12 × r. Thus, in this multiplicative model, translation to 12 hours follows not only the “average” dependence curve but also preserves each patient’s individual level relative to that curve, improving personalization of standardization with a minimal set of input variables.

### Comparison with the eLi_12_ model in five clinical cases (proof of concept)

As shown in Table 4, in a proof-of-concept analysis of five patients with paired lithium concentration measurements, the multiplicative standardization model for the 12-hour level demonstrated slightly higher accuracy than the published eLi_12_ formula. The mean absolute error decreased from 0.056 to 0.042 mEq/L. The improvement was most pronounced in patients with higher lithium concentrations, whereas at lower concentrations and with later blood sampling, variability persisted, reflecting the influence of unmeasured pharmacokinetic factors. These results support the notion that introducing an individual correction factor (the ratio of the measured value to the mean model prediction at the observed sampling time) can partially account for interindividual differences and improve the accuracy of recalculating lithium concentration to the standard 12-hour level. This is because the individual correction factor effectively encodes a patient-specific “excess/deficit” relative to the average curve–potentially reflecting stable interindividual determinants – and carries this adjustment forward to the target 12-hour time point. In other words, the model separates the “average time trajectory” from the “individual deviation from it,” which is clinically closer to personalized standardization.

**Table 4.** Boston proof-of-concept (5 patients from Köhler-Forsberg et al. (2025) [9]): comparison of the published eLi_12_ formula and the multiplicative model.

| Parameters | ID 1 | ID 2 | ID 4 | ID 6 | ID 7 | Mean |
| --- | --- | --- | --- | --- | --- | --- |
| Age range | 18-22 | 23-27 | 28-32 | 43-47 | 23-27 | 29.2 |
| Sex | Female | Male | Male | Male | Male | – |
| Daily lithium dose (mg) | 450 | 900 | 600 | 300 | 750 | 600 |
| Hours after last dose (h) | 2.90 | 4.00 | 3.90 | 4.17 | 6.17 | 4.20 |
| 12-h se-Li (true) | 01.09 | 0.44 | 0.61 | 0.31 | 0.31 | 0.552 |
| 12 + h se-Li (measured) | 0.90 | 0.35 | 0.50 | 0.28 | 0.18 | 0.442 |
| eLi<sub>12</sub> published | 0.97 | 0.41 | 0.57 | 0.34 | 0.25 | 0.508 |
| Deviation eLi<sub>12</sub> (true – pred) | 0.12 | 0.03 | 0.04 | –0.03 | 0.06 | 0.056 |
| **SimpLi** | **1.03** | **0.42** | **0.6** | **0.35** | **0.23** | **0.526** |
| Deviation | 0.06 | 0.02 | 0.01 | –0.04 | 0.08 | 0.042 |
| SimpLi (true – pred) |  |  |  |  |  |  |

## Discussion

In this study, we present and validate a statistical model for recalculating a measured serum lithium concentration to the standard 12-hour level. The results show that the synthetic patient cohort generated from aggregated literature data accurately reproduces key characteristics of real-world samples. Specifically, we were able to replicate the distribution of time between the last dose and blood sampling, the range of lithium doses used and concentrations observed, and the correlational relationships among these parameters. This level of agreement suggests that the simulation model adequately reflects real-world conditions of lithium monitoring and can serve as a reliable basis for evaluating standardization methods.

An assessment of the predictive performance of different recalculation approaches highlighted the limitations of simple linear models. Linear regression using only time since the last dose, although identifying a statistically significant association, explained less than 20% of the variance in the 12-hour level (R^2^ < 0.2). Adding dose as an additional predictor produced only a small increase in explained variance (also R^2^ < 0.2), confirming that a linear relationship based solely on time and dose is insufficient for accurate prediction. In contrast, the multiplicative model incorporating a patient-specific correction factor substantially improved recalculation accuracy. In this approach, the correction factor captures individual features of lithium elimination, resulting in predicted 12-hour concentrations that were much closer to the true values. Introducing an individually adaptable component therefore markedly improved agreement between standardized values and the target level, particularly at higher lithium concentrations, where kinetic variability tends to be more pronounced.

Our recalculation model is aligned with recently proposed methods for standardizing lithium levels. In particular, eLi_12_, introduced by Köhler-Forsberg et al. (2025), is a simple equation for estimating the 12-hour concentration based on the measured level and the elapsed time since the last dose [9]. The authors developed the eLi_12_ formula using clinical trial data and evaluated it in two pilot studies. However, our approach differs in ways that may offer practical advantages. The eLi_12_ method provides a single universal formula for all patients, whereas our multiplicative model includes an individual correction factor that adapts to the characteristics of a specific patient. This may be especially important at the extremes—for example, in patients with higher lithium concentrations or atypical sampling intervals. A universal formula derived from average parameters may perform less accurately at the edges of the distribution, where individual pharmacokinetic curves deviate from the mean. In such situations, personalized recalculation may be more robust. Our findings suggest that incorporating an individual factor improves agreement between the recalculated and the true level, whereas a fixed formula may underestimate concentration in patients with rapid elimination or overestimate it in those with slower elimination. A direct head-to-head comparison of eLi_12_ and our model on the same dataset would be useful to clarify their relative strengths.

Several limitations should be considered when interpreting these findings. First, the proposed model does not include explicit pharmacokinetic parameters, such as clearance or volume of distribution. In essence, recalculation relies on statistical relationships rather than a full physiological description of lithium kinetics. This means that the model does not directly account for individual differences in renal function or tissue distribution. In real populations, variability in these parameters can be substantial, and without their explicit inclusion the model may lose accuracy in atypical cases.

Second, the simulation was performed only once (nsim = 1), meaning that all conclusions are based on a single generated dataset. Although the dataset was constructed using distributions that approximate real-world data, the lack of repeated simulations prevents quantification of the variability of model results under stochastic fluctuations. In other words, we did not conduct a formal sensitivity analysis of model performance to random population variability. The absence of repeated verification introduces some uncertainty regarding the generalizability of the reported statistical estimates: their confidence intervals may be wider than they appear and require further refinement.

Another limitation is that the model did not account for adherence-related and clinical-state factors that can influence lithium levels. We assumed strict adherence to dosing (taken daily at the same time, with no missed doses) and clinical stability (no acute changes in hydration, comorbid conditions, etc.). In real-world practice, these assumptions are often violated. For example, missed or delayed doses, intentional regimen changes by patients, or acute dehydration episodes (e.g., due to hot weather, vomiting, or diarrhea) can markedly alter serum lithium levels independently of time since the last dose. Nonadherence remains a major challenge in lithium treatment; while a recalculation model can correct for sampling-time error, it cannot compensate for errors caused by incorrect medication intake. Future work should incorporate these realities, either by extending the model or by more clearly delineating the conditions under which it is applicable.

The present findings suggest several directions for further development and implementation of a 12-hour lithium standardization model. First, integrating the proposed standardization into clinical decision support systems appears promising. In modern electronic health systems, an automated recalculation could be implemented such that, alongside the laboratory-reported lithium concentration, an equivalent 12-hour value would be provided based on the documented time of the last dose. This functionality would substantially facilitate clinical workflows by allowing clinicians to interpret results immediately in the context of the therapeutic range. Even if a patient’s blood draw did not occur exactly 12 hours post-dose, the clinician would still see a corrected value that is comparable with the target corridor (e.g., 0.6–0.8 mmol/L for relapse prevention). As shown by Köhler-Forsberg et al., allowing flexibility in sampling time while applying mathematical correction can improve convenience without compromising monitoring accuracy [9]. Ultimately, embedding such recalculation functionality into decision support systems may increase adherence to regular lithium monitoring (given that approximately half of samples are currently obtained outside the recommended window) and thereby improve overall quality of bipolar disorder care.

The next step should be to test the model using real clinical data. While a synthetic cohort provides initial validation, definitive confirmation of utility requires evaluation in patient samples. This could be done retrospectively, for example using data from patients who, for independent reasons, had lithium measured at different time points. An ideal scenario would be a prospective validation: in a patient group, lithium could be measured at two different times (e.g., at a nonstandard time and again at 12 hours), and the recalculated estimates could be compared against the observed 12-hour level. Such a study would allow direct assessment of accuracy and clinical usefulness in a real population and would enable comparison with alternative approaches (including the eLi_12_ equation) under routine practice conditions. In addition, real-world data may reveal systematic biases that are difficult to anticipate in simulations.

## Conclusion

This study demonstrates the feasibility and practical value of using a statistical model (SimpLi) to standardize a measured serum lithium concentration to an estimated 12-hour level. The synthetic cohort, constructed from aggregated data from real studies, reproduced key parameters with high accuracy – including the distributions of blood sampling times, doses, concentrations, and their interrelationships – supporting the adequacy of the chosen approach for modeling this problem.

The proposed multiplicative model incorporating an individual correction factor outperformed simple linear regressions based only on time and dose. Its main advantage is the ability to preserve a patient’s individual deviation from the average population trajectory, enabling a more personalized adjustment – particularly relevant at extreme concentration values or atypical sampling times. In a small pilot comparison with the existing eLi_12_ method, the results suggested improved accuracy, with the mean absolute deviation from the true level decreasing from 0.056 to 0.042 mmol/L.

Overall, SimpLi is a grounded tool for correcting lithium therapeutic drug monitoring results when the standard 12-hour sampling interval is not met. Potential implementation in laboratory information systems and electronic health records as an automated function could reduce clinical misinterpretation, improve treatment safety, and promote more effective use of real-world data. Before clinical deployment, further validation is required using independent real-world datasets with precise documentation of dosing and blood sampling times.

## Data Availability

All materials required to reproduce the simulation and analyses (code and synthetic dataset) are publicly available via Zenodo. The web implementation of SimpLi is available online.
Zenodo (code + synthetic dataset): https://zenodo.org/records/18380482
SimpLi web tool: https://psyandneuro.ru/bekhterev-ai/simpli/

https://psyandneuro.ru/bekhterev-ai/simpli/

## Additional information

### Author contributions (CRediT)

Evgeny Kasyanov: methodology; software; formal analysis; data curation (synthetic data generation); visualization; writing-original draft; writing-review & editing. Galina E. Mazo: conceptualization; supervision; writing-review & editing.

### Ethics approval and consent to participate

This study used exclusively synthetic (simulated) data and did not involve human participants, identifiable personal data, or biological materials. Therefore, ethics committee approval and informed consent were not required.

### Consent for publication

Not applicable.

### Manuscript status (submission statement)

This manuscript has **not** been submitted to any journal and is not under consideration elsewhere.

### Data and code availability

All materials required to reproduce the simulation and analyses (code and synthetic dataset) are publicly available via Zenodo. The web implementation of SimpLi is available online.

Zenodo (code + synthetic dataset): https://zenodo.org/records/18380482

SimpLi web tool: https://psyandneuro.ru/bekhterev-ai/simpli/

### Use of generative AI

Generative AI was used to assist with partial R code development and editing of manuscript text. All AI-assisted outputs were reviewed and edited by the authors, who take full responsibility for the content of the manuscript, including its accuracy, integrity, and compliance with publication standards.

